# Paired plaque and plasma proteomics reveal molecular signatures of symptomatic atherosclerosis

**DOI:** 10.64898/2026.08.23.26361143

**Authors:** Lanyue Zhang, Luka Živković, Anushree Ray, Roya Batool, Julian Louma, Iulia Lupul, Mohamad Ali Antabi, LingLing Xu, Paulo Vinicius Gil Alabarse, Jan Stana, Abdalla Marei, Nikolaos Tsilimparis, Marios K. Georgakis

## Abstract

**Background:** Phenotyping of atherosclerotic plaque vulnerability has largely relied on histopathology that captures structural features, but does not fully account for clinical presentation. Proteomic profiling could uncover molecular readouts of vulnerability that refine plaque phenotyping and provide mechanistic insights. Yet, the proteomic signatures associated with plaque rupture and symptomatic presentation are poorly characterized.

**Methods:** We profiled paired carotid plaque tissue and preoperative plasma from 88 patients undergoing carotid endarterectomy (51 symptomatic, 37 asymptomatic) using the Olink Explore 3072 platform. We related plaque protein abundance to symptomatic presentation and quantitative histopathological features, and compared the performance of histopathology-vs. proteomics-based models for discriminating symptomatic disease. Next, we developed proteomic signatures of cellular abundance and explored their associations with plaque phenotypes by using plaque single-cell RNA-sequencing (scRNA-seq) data. Finally, we assessed plaque–plasma concordance across 2,837 shared proteins.

**Results:** Across 2,837 plaque proteins, 19 were differentially expressed in symptomatic plaques related to distinct clinical events, highlighting pathways related to neutrophil degranulation and innate immune system. FGFBP1 showed the strongest association with symptomatic presentation (log_2_ fold change = 1.14; P = 1.82 × 10^-6^). Proteins associated with a composite vulnerability index based on histopathology were enriched for inflammatory pathways, including TNFα signaling through NFκB, complement activation, and IL6–JAK–STAT3 signaling. Individual proteins also mapped to specific histopathological features, including CXCL8 associated with macrophage burden and lipid core size, and EPHB4 and PKN3 with neovascularization. A proteomics-based model discriminated symptomatic from asymptomatic plaques substantially better than a histopathology-based model (AUC 0.83 vs. 0.66; P = 0.026). Integration with scRNA-seq data enabled the development of cell-class signatures that correlated with histopathology readouts, including macrophage burden, smooth muscle cell content, and neovascularization. Plaque and plasma protein levels showed limited overall correspondence (median ρ=0.11), although selected proteins, including FGFBP1, demonstrated concordant associations in plasma.

**Conclusions:** Deep proteomic profiling of human carotid plaques identifies molecular signatures of symptomatic atherosclerosis that extend beyond conventional histopathology. These signatures implicate neutrophil activation and inflammatory signaling pathways as key determinants of plaque vulnerability. Although plaque and plasma proteomes are largely distinct, selected proteins may represent promising circulating biomarkers for future risk stratification.

## Introduction

Cardiovascular disease remains the leading cause of morbidity and mortality worldwide.^1,2^ The rupture of atherosclerotic plaques underlies the majority of acute symptomatic cardiovascular events, such as myocardial infarction and ischemic stroke.^3,4^ Histopathological features such as macrophage infiltration, lipid core, intraplaque hemorrhage, calcification, and fibrous cap status form the basis of the current conceptual framework of plaque vulnerability.^5^ Yet these features may also be present in asymptomatic plaques and are not uniformly observed in symptomatic lesions.^6–8^ These observations suggest that plaque morphology incompletely explains why some lesions become clinically symptomatic.

Proteomic profiling can provide a molecular view of human plaque biology and can reveal disease-associated features that are not directly captured by conventional histopathology.^9–11^ Identifying proteomic signatures of plaque vulnerability could improve risk stratification of plaques, refine plaque phenotyping in research settings, and offer mechanistic insights with implications for drug development. However, the proteomic phenotypes associated with plaque vulnerability remain incompletely characterized. Previous human plaque proteomic studies have generally involved modest sample sizes,^10,12,13^ relied on restricted proteomic coverage,^10,14^ focused on individual morphological endpoints,^15–17^ and explored specific plaque areas^18^ or biological mechanisms.^19^ Although larger studies have integrated plaque proteomics with histopathological and clinical phenotyping,^9,18,19^ the relationship between symptom-associated and histopathology-associated proteomic signals remains incompletely defined. Whether plaque proteomics provides information beyond histopathology for symptomatic presentation has not been directly tested. Furthermore, only a limited number of studies have allowed comparisons with other plaque omics layers^15,16,19^ or performed paired analyses with plasma proteomics in the same patients.^14,16^

To address these gaps, we used the Olink Explore 3072 platform to profile paired carotid plaque and preoperative plasma proteomes from 88 patients undergoing carotid endarterectomy in the AtherOMICS Biobank.^20^ We characterized plaque protein programs associated with symptomatic presentation and clinically defined ischemic events. We then related plaque proteins to quantitative histopathological features, and tested whether proteomic measurements added discriminative information beyond histology. We integrated these data with human carotid plaque single-cell RNA sequencing (scRNA-seq) to develop proteomic signatures of the cellular composition of human plaques. Finally, we examined plaque–plasma correspondence to explore whether lesion-associated molecular signals are detectable in the circulation.

## Methods

### Study design

Participants were drawn from the AtherOMICS biobank, an ongoing study at LMU Klinikum (Munich, Germany) enrolling patients scheduled for carotid or femoral endarterectomy since August 2022. AtherOMICS systematically collects paired plaque tissue and peripheral blood from the same patient, alongside *in vivo* plaque imaging and clinical data, to enable integrated multi-omic profiling. Details on the study protocol have been described previously.^20^

By September 2024, the AtherOMICS biobank had enrolled 154 patients. The present analysis comprised the first 88 consecutive eligible patients prioritized for Olink Explore 3072 proteomic profiling based on availability of matched pre-operative plasma, serum, and carotid plaque tissue, as well as linked histology and clinical data. Of these, 51 had symptomatic carotid stenosis defined by ipsilateral stroke, transient ischemic attack, amaurosis fugax, or central retinal artery occlusion attributable to the operated lesion. The remaining 37 had asymptomatic carotid stenosis. Indication for carotid endarterectomy was made by the treating clinical team according to standard institutional practice. Matched single-nucleus RNA-sequencing (snRNA-seq) data were available for 15 of these patients.

The study was approved by the Ethics Committee of the LMU Faculty of Medicine (approval numbers 121-09, 22-0135, 23-0772). All participants provided written informed consent.

### Sample collection and processing

Carotid plaques were retrieved from the operating room and transferred to the laboratory on ice in autoclaved phosphate-buffered saline. The median total processing time from surgical removal to flash-freezing was 30 minutes and did not exceed 60 minutes. Each plaque was sectioned into 5 mm segments, with the most-diseased segment (MDS) in the internal carotid artery and adjacent 5 mm segments proximal and distal to it identified by morphological assessment. The MDS was reserved for histopathology after fixation in 4% paraformaldehyde. Adjacent segments were flash-frozen in 2-methylbutane pre-cooled in liquid nitrogen and stored at −80 °C for downstream proteomic and transcriptomic analyses. Peripheral blood was collected into 9-mL EDTA tubes before surgery. After 30 minutes at room temperature, plasma was separated by centrifugation at 2,000 × g for 10 minutes at 15 °C, aliquoted into cryovials at 300 µL per aliquot, and stored at −80 °C until proteomic profiling.

### Plaque and plasma proteomic profiling

For plaque proteomic profiling, frozen plaque segments were homogenized and lysed in RIPA buffer supplemented with complete protease inhibitor cocktail (Sigma-Aldrich), filtered through 70 µm cell strainers, and centrifuged at 14,000 rpm at 4 °C to remove debris. Sample integrity was assessed by SDS-PAGE with Coomassie Blue staining prior to proteomic profiling. Plaque lysates and matched plasma samples from the same patients were loaded onto 96-well plates and processed at the Metabolomics and Proteomics Core, Helmholtz Zentrum München (Munich, Germany). Proteomic profiling was conducted using the antibody-based Olink Explore 3072 proximity extension assay (PEA) across eight 384-plex panels (cardiometabolic, cardiometabolic II, inflammation, inflammation II, neurology, neurology II, oncology, and oncology II). Analyses were based on Normalized Protein eXpression (NPX) values after quality control. Samples and assays receiving a QC warning in the Olink output were excluded from downstream analyses.

### Plaque histopathology

The MDS was decalcified, dehydrated, and embedded in paraffin. Serial sections of 5 µm thickness were taken at 1 mm intervals and stained with hematoxylin and eosin, Picrosirius red, anti-CD68 immunohistochemistry, and anti-α-smooth muscle actin (αSMA) immunohistochemistry. Whole-slide images were acquired with a high-resolution slide scanner (Carl Zeiss) and analyzed in QuPath (v0.4.4). CD68-positive area and αSMA-positive area were quantified using QuPath’s random trees pixel classifier trained on slide-specific annotations. Lipid core, intraplaque hemorrhage, calcification, neovascularization (represented as percentages of total plaque area), plaque area, and fibrous cap thickness were manually annotated on the whole-slide images by experienced staff blinded to clinical and proteomic data, with divergent annotations resolved by consensus. Fibrous cap rupture and erosion were identified by visual assessment and recorded as binary variables. We additionally constructed a composite plaque vulnerability index (PVI, range 0–4), adapted from a previously described histological vulnerability index,^21^ with one point assigned for heavy macrophage infiltration (CD68-positive area), low smooth muscle cell (SMC) content (αSMA-positive area), large lipid core, or extensive intraplaque hemorrhage, each defined as the upper or lower cohort-internal quartile.

### Tracing cellular origin of plaque proteins

To trace the cellular origin of plaque proteins, we applied an approach adapted from Tracing Expression of Multiple Protein Origins^22^ (TEMPO) to a human plaque scRNA-seq reference atlas by integrating four publicly available human carotid plaque datasets comprising 36 donors (16 symptomatic and 20 asymptomatic).^23–26^ The datasets were processed using a uniform Seurat-based workflow, including quality control, SCTransform normalization, doublet removal, and principal component analysis. Dataset integration and batch correction were performed using Harmony (v1.2.3),^27^ accounting for sample identity and sequencing technology, followed by graph-based clustering of the integrated embedding. Cell types were annotated using canonical marker genes obtained from published single-cell studies and atlases of human atherosclerotic and vascular tissues.^28^

For each gene, average normalized expression for each major cell class was calculated from donor/sample-level pseudobulk profiles within each dataset and then averaged across datasets before z-scoring across cell classes, such that the z-score represented relative enrichment of that gene in one plaque cell class compared with the other plaque cell classes. The class with the highest z-score was designated as the candidate source cell class. Genes were retained if they met three criteria in their candidate cell class: a z-score ≥ 2 in the assigned class, average normalized expression above the 10th percentile of all detected genes, and expression at least two-fold higher than in the second-highest cell class. Source genes were linked to Olink assays through UniProt identifiers, and only assays with median plaque NPX above the 10th percentile across the cohort were retained to reduce the influence of low-abundance assays. For each major cell class, a sample-level proteomic signature was then computed as the first principal component (PC1) of standardized NPX values across the assigned assays.

To examine whether the proteomic signatures captured cellular biology, signatures were correlated with histopathological measurements using Spearman correlation. In the 15 plaques with paired snRNA-seq data, signatures were additionally compared with the corresponding snRNA-seq-derived cell-class fraction. snRNA-seq processing followed the AtherOMICS protocol.^20^

### Statistical analysis

Continuous variables were summarized and compared according to their distribution: approximately symmetric variables as mean (SD) using t-tests, and skewed variables as median (IQR) using Wilcoxon rank-sum tests. Categorical variables were summarized as n (%) and compared using χ² tests. Missing histopathological measurements were handled using multiple imputation by chained equations with classification and regression trees in the *mice (3.19.0)* R package (m = 50, maxit = 20). Imputed estimates were pooled using Rubin’s rules unless otherwise stated, and complete-case analyses were performed as sensitivity analyses.

#### Proteome variability analysis

To assess the contribution of age, sex, and symptomatic status to protein-level variability, we performed protein-wise Type III ANOVA separately in plaque and plasma using the *car (3.1.3)* R package. The variance explained by each factor was expressed as η² with the *effectsize (1.0.1)* package and visualized as stacked variance decomposition plots.

#### Differential protein abundance analysis

To identify proteins associated with symptomatic status, linear models implemented in *limma (3.64.3)* with robust empirical Bayes moderation were fitted separately to plaque and plasma proteomics data, adjusting for age and sex. Comparisons included symptomatic versus asymptomatic carotid stenosis as the primary contrast and three clinically stratified contrasts: Stroke/Retinal Artery Occlusion (RAO) versus asymptomatic, symptomatic non-stroke (transient ischemic attack [TIA], amaurosis fugax, and other) versus asymptomatic, and Stroke/RAO versus symptomatic non-stroke.

#### Histopathology-proteome association analysis

Associations of histopathological features and the plaque vulnerability index with plaque protein abundance were assessed using age- and sex-adjusted protein-wise linear models. Models were fitted separately for each histopathological feature or index. Pathology features with right-skewed distributions were log1p-transformed before z-score standardization. For PVI, protein abundance was modelled as the outcome and the ordered PVI score (0–4) was used as the predictor to test for trends across increasing plaque vulnerability.

#### Gene set enrichment analysis (GSEA)

Preranked GSEA was performed using the t statistics from the corresponding association tests, querying the Reactome database via the *ReactomePA (1.52.0)* R package for the differential protein abundance analyses and the Molecular Signatures Database (MSigDB) Hallmark gene sets for the plaque vulnerability index analyses.

#### Correlation analysis

Age- and sex-adjusted partial Spearman correlations were used to assess pairwise relationships among continuous histopathological features and within-protein plaque–plasma correlations for proteins measured in both compartments. Correlations involving imputed histopathological data were estimated within each imputed dataset and pooled on the Fisher z scale.

#### Association and discrimination analysis for symptomatic status

To evaluate the relationship of plaque features with symptomatic status, individual histopathological features were first tested using logistic regression models adjusted for age and sex. Odds ratios for continuous features are reported per 1-SD increase, and fibrous cap rupture and erosion were modelled as binary variables.

Apparent discriminative performance for symptomatic status was then assessed using receiver operating characteristic area under the curve (AUC). Single-feature apparent AUCs were calculated for individual histopathological features and plaque proteins, with 95% confidence intervals estimated using DeLong’s method. All multivariable logistic regression models were adjusted for age and sex. We compared a baseline model with age and sex alone, a model adding intraplaque hemorrhage and CD68-positive area, a model adding the top three plaque proteins, and a combined model with both. For analyses involving imputed histopathological features, AUC estimates and paired DeLong comparisons between models were pooled across imputations using Rubin’s rules. ROC curves were drawn from sample-level mean predicted probabilities across imputations.

All analyses were performed in R version 4.5.1. All P values were two-sided. For analyses involving multiple hypothesis testing, P values were adjusted using the Benjamini-Hochberg false discovery rate (FDR).

## Results

### Summary of study design

We profiled paired plaque tissue and pre-operative plasma samples from 88 patients undergoing carotid endarterectomy using the Olink Explore 3072 platform, enabling within-patient comparison of the lesion and circulating proteomes. Plaque tissue was additionally characterized by quantitative histology and matched single-nucleus RNA sequencing data that were available for a subset of the patients (n=15). The overall study design and analytical workflow are summarized in **Figure 1**. Of the 88 patients, 51 were symptomatic and 37 asymptomatic carotid stenosis (mean age 72.1 years, 30.7% female; **Table 1**). The two groups were broadly comparable with regard to age and sex. Symptomatic patients more frequently were current smokers and had higher LDL and total cholesterol, while the remaining characteristics did not differ significantly between groups (**Table 1**).

**Figure 1.**
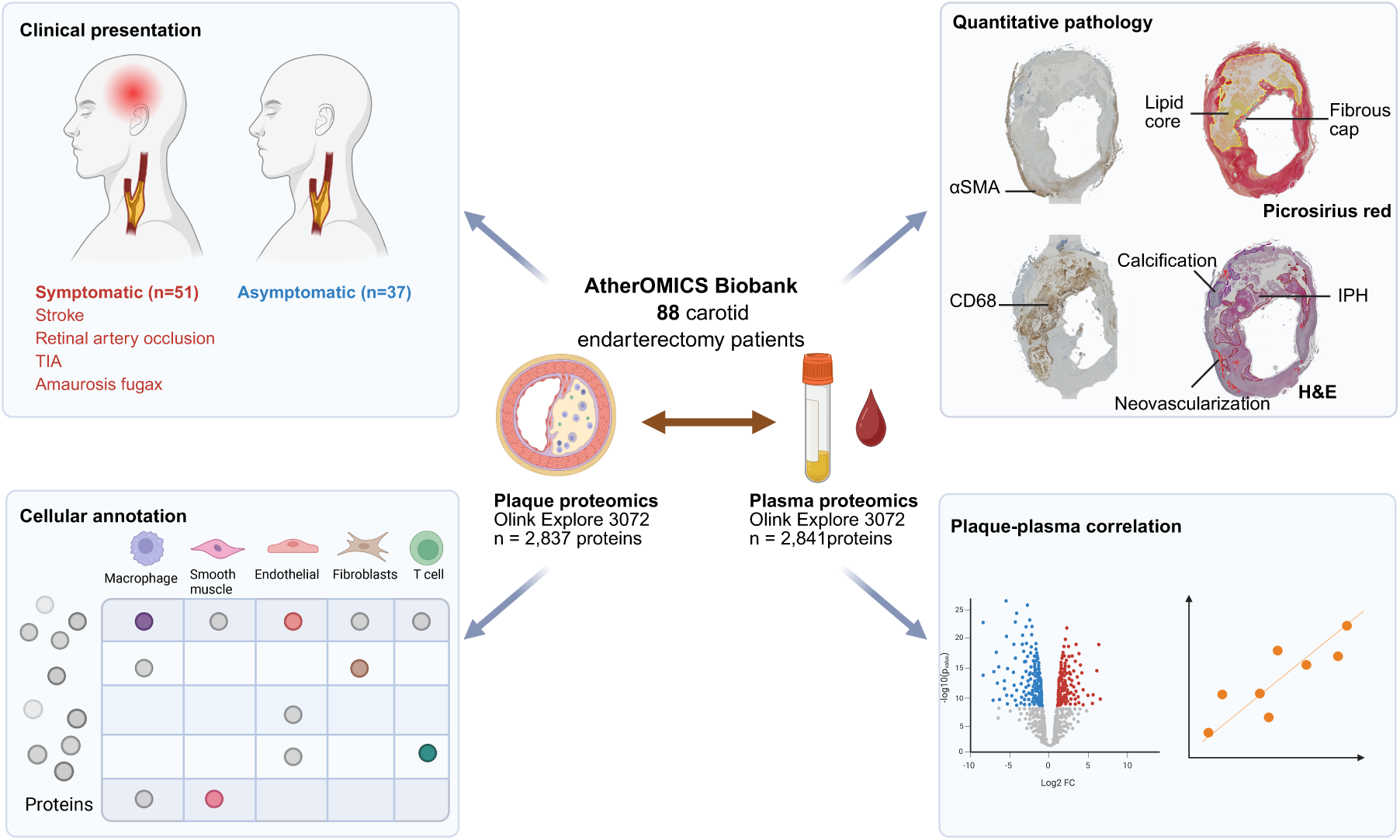
Study design. The study included 88 patients undergoing carotid endarterectomy with paired plaque tissue and pre-operative plasma. Plaque tissue was profiled by Olink Explore 3072 proteomics and quantitative histopathology, and matched plasma underwent paired proteomic profiling. Human carotid plaque single-cell references and a matched single-nucleus RNA-sequencing subset were used to trace candidate cellular origins of plaque protein signals. Downstream analyses examined plaque proteins associated with clinical presentation, quantitative histopathological features, incremental symptom-discriminative information beyond histology, candidate cellular origins, and plaque–plasma concordance.

**Table 1.** Clinical characteristics of AtherOMICS patients with available paired plaque and plasma proteomics data.

| **Variable** | **Overall**<br>**N = 88** | **Asymptomatic**<br>**N = 37** | **Symptomatic**<br>**N = 51** | **P-value** |
| --- | --- | --- | --- | --- |
| **Sex, n (%)** |  |  |  | 0.527 |
| Female | 27 (30.7%) | 10 (27.0%) | 17 (33.3%) |  |
| Male | 61 (69.3%) | 27 (73.0%) | 34 (66.7%) |  |
| **Age (years), mean (SD)** | 72.1 (8.3) | 72.0 (7.1) | 72.3 (9.1) | 0.871 |
| **BMI (kg/m<sup>2</sup>), median (IQR)** | 25.7 (23.2, 28.1) | 25.8 (23.4, 27.9) | 25.4 (23.0, 29.1) | 0.87 |
| Unknown | 1 | 1 | 0 |  |
| **Diabetes mellitus, n (%)** | 23 (26.7%) | 7 (19.4%) | 16 (32.0%) | 0.194 |
| Unknown | 2 | 1 | 1 |  |
| **Smoking status, n (%)** |  |  |  | **0.031** |
| No | 27 (33.3%) | 13 (41.9%) | 14 (28.0%) |  |
| Yes, former | 33 (40.7%) | 15 (48.4%) | 18 (36.0%) |  |
| Yes, current | 21 (25.9%) | 3 (9.7%) | 18 (36.0%) |  |
| Unknown | 7 | 6 | 1 |  |
| **Statin use, n (%)** | 66 (75.0%) | 30 (81.1%) | 36 (70.6%) | 0.262 |
| **LDL cholesterol (mg/dL), mean (SD)** | 94.0 (41.7) | 77.8 (36.8) | 104.7 (41.6) | **0.004** |
| Unknown | 10 | 6 | 4 |  |
| **HDL cholesterol (mg/dL), mean (SD)** | 49.6 (13.4) | 53.2 (15.4) | 47.1 (11.4) | 0.066 |
| Unknown | 12 | 6 | 6 |  |
| **Total cholesterol (mg/dL), mean (SD)** | 159.1 (42.5) | 143.5 (35.2) | 169.6 (44.0) | **0.005** |
| Unknown | 11 | 6 | 5 |  |
| **HbA1c (%), median (IQR)** | 6.0 (5.6, 6.4) | 5.8 (5.5, 6.2) | 6.0 (5.6, 6.6) | 0.103 |
| Unknown | 16 | 8 | 8 |  |
| **C-reactive protein (mg/dL), median (IQR)** | 0.3 (0.1, 0.5) | 0.1 (0.1, 0.5) | 0.3 (0.1, 0.6) | 0.065 |
| Unknown | 1 | 0 | 1 |  |
| **Hypertension, n (%)** | 72 (82.8%) | 33 (91.7%) | 39 (76.5%) | 0.065 |
| Unknown | 1 | 1 | 0 |  |
| **Coronary heart disease, n (%)** | 33 (38.8%) | 15 (41.7%) | 18 (36.7%) | 0.645 |
| Unknown | 3 | 1 | 2 |  |
| **Prior stroke, n (%)** | 21 (30.0%) | 6 (20.7%) | 15 (36.6%) | 0.153 |
| Unknown | 18 | 8 | 10 |  |
| **Peripheral arterial disease, n (%)** | 19 (23.2%) | 7 (20.0%) | 12 (25.5%) | 0.557 |
| Unknown | 6 | 2 | 4 |  |
| **Family history of CVD, n (%)** | 45 (60.0%) | 20 (64.5%) | 25 (56.8%) | 0.503 |
| Unknown | 13 | 6 | 7 |  |

n (%); Mean (SD); Median (Q1, Q3)
Pearson's Chi-squared test; Welch Two Sample t-test; Wilcoxon rank sum test
Abbreviation: BMI, body mass index; LDL-C, low density lipoprotein cholesterol; HDL-C, high density lipoprotein cholesterol; HbA1c, glycated hemoglobin A1c.

### A focused plaque proteomic signal is associated with symptomatic presentation

To assess whether the plaque proteome differs by clinical presentation, we first partitioned protein-level variance across the 2,837 quantified plaque proteins for age, sex, and symptomatic status. Across the plaque proteome, these factors explained only a small fraction of the variance, with median values below 1% for each (**Figure 2A, Supplementary Table 1**). Nevertheless, individual proteins showed stronger associations. The top-ranked proteins were the prostate-specific antigen KLK3 for sex (η² = 51.3%), CRYBB2 for age (η² = 22.7%), and FGFBP1 for symptomatic status (η² = 23.4%; **Figure 2B, Supplementary Table 1**).

**Figure 2.**
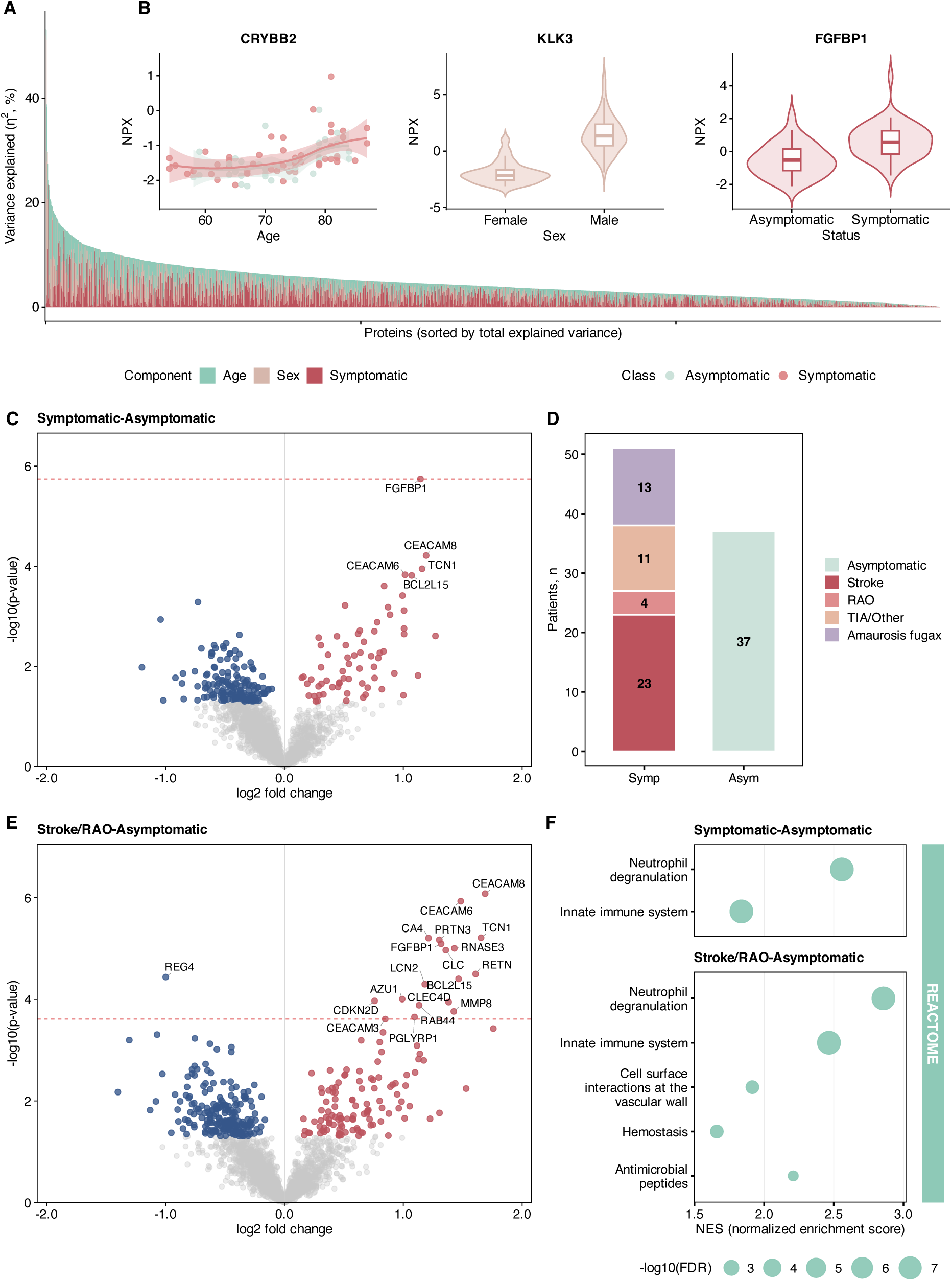
Plaque proteomics implies plaque biology related to symptomatic atherosclerosis. **(A)** Contribution of age, sex, and symptomatic status to plaque proteome variability. For each protein, a linear model including age, sex, and symptomatic status was fitted, and Type III ANOVA was used to estimate the proportion of variance explained by each model term (η²). Proteins are ordered by total explained variance. **(B)** Representative proteins showing the strongest associations with symptomatic status (FGFBP1), sex (KLK3), and age (CRYBB2). **(C)** Symptomatic versus asymptomatic plaque proteome comparison. Volcano plots were generated from age- and sex-adjusted limma models. The x-axis shows log2 fold change and the y-axis shows −log10 (raw P value). Proteins with nominal P < 0.05 are colored, and the dashed red line marks the raw-P threshold corresponding to FDR = 0.05. Labels indicate proteins with FDR < 0.10. **(D)** Distribution of clinical presentation in the analyzed cohort. Numbers within bars indicate patient counts. Symptomatic presentations included stroke, TIA, amaurosis fugax, RAO, and other/unknown manifestations. **(E)** Stroke/RAO versus asymptomatic plaque proteome comparison. Volcano plots were generated from direct pairwise age- and sex-adjusted limma models. The x-axis shows log2 fold change and the y-axis shows −log10(raw P value). Proteins with nominal P < 0.05 are colored, and the dashed red line marks the raw-P threshold corresponding to FDR = 0.05. Labels indicate proteins with FDR < 0.05. **(F)** Reactome gene set enrichment analysis based on ranked differential abundance results for the symptomatic– asymptomatic and stroke/RAO–asymptomatic comparisons. Bubble size indicates enrichment significance [−log10(FDR)], and the x-axis shows the normalized enrichment score (NES). Abbreviations: FDR, false discovery rate; TIA, transient ischemic attack; RAO, retinal artery occlusion; NPX, normalized protein expression.

We next compared protein abundance between symptomatic and asymptomatic plaques. Although 226 proteins differed at nominal significance, only FGFBP1 remained significant after correction for multiple testing, with higher abundance in symptomatic plaques (log2FC = 1.14, P = 1.82 × 10^-6^, FDR = 0.0052; **Figure 2C, Supplementary Table 2**). We hypothesized that the small number of differentially abundant proteins between symptomatic and asymptomatic plaques might reflect the clinical heterogeneity of the symptomatic group, which included stroke (n=23), RAO (n=4), TIA/other (n=11), and amaurosis fugax (n=13) (**Figure 2D**). We therefore focused on plaques from patients with stroke or RAO that represent ischemic events with objective imaging correlates (**Figure 2E, Supplementary Table 3**). This sharper contrast strengthened the proteomic signal and identified 19 proteins at FDR < 0.05, with FGFBP1 remaining among the leading proteins (log2FC = 1.32, P = 8.06 × 10^-6^, FDR = 0.0038), alongside neutrophil activation and granule-associated markers such as CEACAM8, PRTN3, and AZU1. Reactome enrichment further highlighted neutrophil degranulation and innate immune pathways (**Figure 2F, Supplementary Table 4**). These analyses indicate that symptomatic carotid plaques carry a focused proteomic phenotype that points to innate immune and neutrophil-associated biology.

### Plaque proteomics provides molecular readouts of histopathology and plaque vulnerability

We next asked whether the plaque proteome reflected plaque vulnerability as captured by quantitative histopathological features. We related plaque protein abundance to seven histological features in age- and sex-adjusted models: CD68-positive area, lipid core, αSMA-positive area, fibrous cap thickness, intraplaque hemorrhage, calcification, and neovascularization, displaying proteins significant (FDR < 0.05) for at least one feature (**Figure 3A, Supplementary Table 5**). Significant associations were concentrated in lipid core, CD68-positive area, calcification, and neovascularization, whereas fibrous cap thickness, intraplaque hemorrhage, and αSMA-positive area did not yield FDR-significant proteins. C-X-C motif chemokine ligand 8 (CXCL8) was the dominant signal among the inflammatory features, increasing with both lipid core size (β = 0.97, FDR = 9.0 × 10^-5^) and CD68-positive area (β = 0.90, FDR = 9.5 × 10^-4^), consistent with its role in a macrophage-neutrophil crosstalk chemokine.^29^ This neutrophil biology also linked symptomatic presentation to plaque histopathology measurements. Among the stroke/RAO–associated proteins, the 13 mapping to neutrophil degranulation were positively associated with CD68-positive area and intraplaque hemorrhage, and negatively with αSMA-positive area, with nominal significance for CD68-positive area in 11 of 13, intraplaque hemorrhage in 8, and αSMA-positive area in 6 (**Supplementary Table 5**). Neovascularization showed the largest number of associated proteins, including EPH receptor B4 (EPHB4; β = 0.28, FDR = 0.010) and protein kinase N3 (PKN3; β = 0.65, FDR = 0.010), both previously linked to angiogenic biology,^30,31^ as well as double C2 domain beta (DOC2B; β = 0.60, FDR = 0.010). Calcification-associated proteins showed predominantly inverse associations between protein levels and calcification burden (**Figure 3A, Supplementary Table 5**). Complete-case sensitivity analyses yielded similar patterns (**Supplementary Figure 1**).

**Figure 3.**
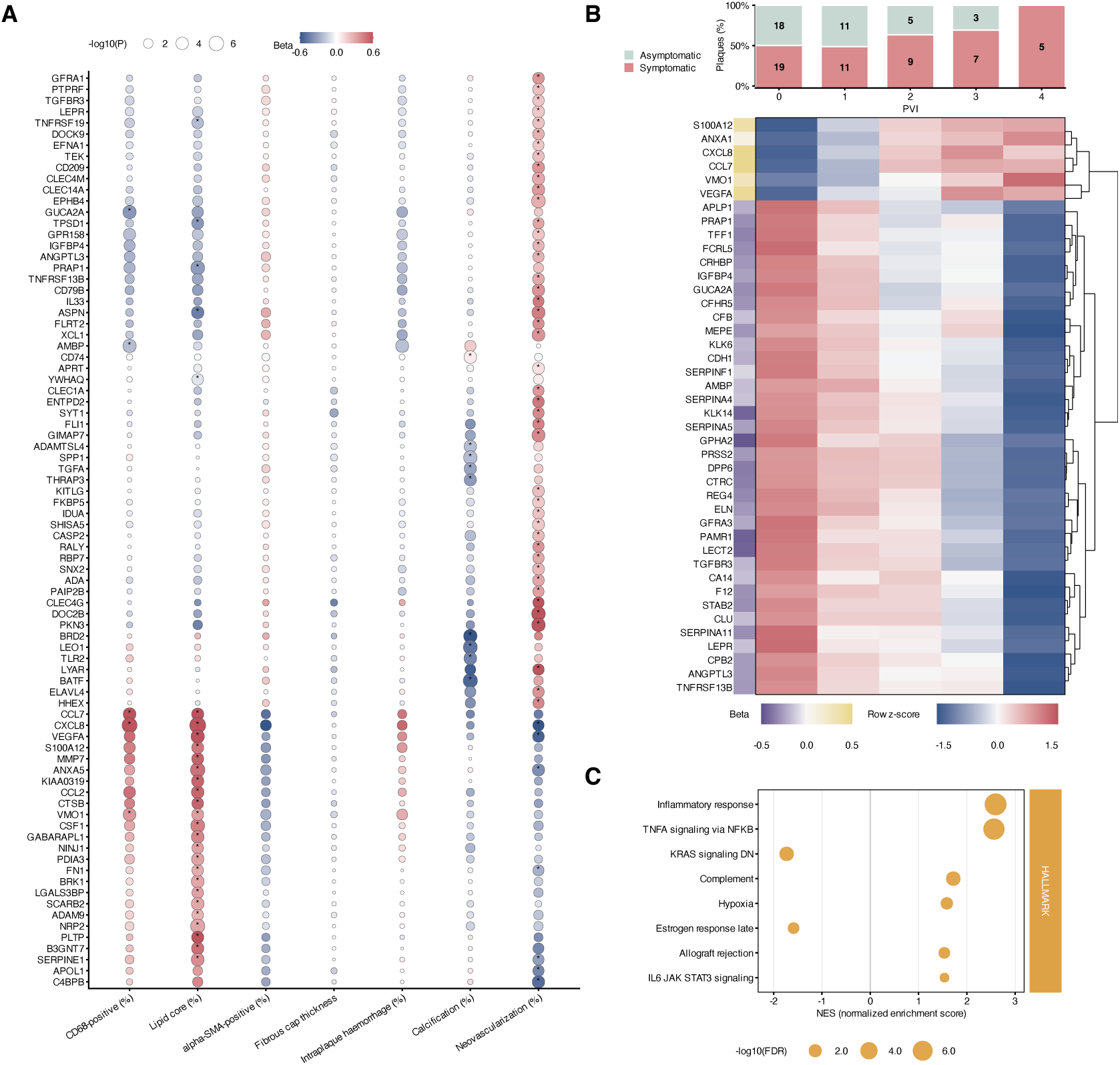
Plaque proteomics maps molecular programs across quantitative histopathological features. Using the imputed pathology data, plaque protein abundance was analyzed in relation to histopathological featur es and the plaque vulnerability index (PVI). **(A)** Bubble plot showing the union of plaque proteins significantly associated with individual histopathological features in feature-specific regression analyses. Columns represent CD68-positive area, lipid core, αSMA-positive area, fibrous cap thickness, intraplaque hemorrhage, calcification, and neovascularization. Bubble color indicates the regression coefficient (beta), bubble size indicates -log10(P value), and asterisks denote FDR < 0.05. **(B)** Heatmap of plaque proteins significantly associated with the PVI. Columns represent PVI groups from 0 to 4, and values are shown as row z-scores of age- and sex-adjusted mean protein abundance. The side annotation shows the PVI association coefficient. Stacked bars above the heatmap show the proportions of symptomatic and asymptomatic plaques within each PVI group, with numbers indicating patient counts. **(C)** Hallmark pathway enrichment analysis based on proteins ranked by their PVI association statistics. Bubble position indicates the normalized enrichment score (NES), and bubble size indicates -log10(FDR).

To summarize the combined burden of established vulnerability features, we further constructed a composite PVI (range 0–4), integrating high macrophage burden, low smooth muscle content, large lipid core, and extensive intraplaque hemorrhage. The proportion of symptomatic plaques increased with PVI, from 51.4% (19/37) plaques at PVI 0 to 100% (5/5) at PVI 4 (**Figure 3B, Supplementary Table 5**). Across the 42 proteins associated with PVI at FDR < 0.05, most decreased toward higher PVI, including proline rich acidic protein 1 (PRAP1; β = −0.30, FDR = 5.9 × 10^-4^), carboxypeptidase B2 (CPB2; β = −0.27, FDR = 2.5 × 10^-3^), and peptidase domain containing associated with muscle regeneration 1 (PAMR1; β = −0.44, FDR = 2.5 × 10^-3^), while CXCL8 (β = 0.73, FDR = 8.5 × 10^-4^), C-C motif chemokine ligand 7 (CCL7; β = 0.65, FDR = 9.5 × 10^-3^), and S100 calcium binding protein A12 (S100A12; β = 0.38, FDR = 2.4 × 10^-2^) increased (**Figure 3B, Supplementary Table 5**). Hallmark pathway enrichment of PVI-ranked proteins highlighted inflammatory response and TNFα signaling via NFκB, together with complement, hypoxia, and IL-6–JAK–STAT3 signaling (**Figure 3C, Supplementary Table 6**). Taken together, these findings reveal a proinflammatory and proangiogenic shift at the proteome level that underlies morphological plaque vulnerability.

### Plaque proteomics adds information for discriminating symptomatic presentation beyond measured histopathology

Although the stroke/RAO-associated neutrophil-related proteins showed concordant associations with plaque histopathology, the protein set which differentiates clinical presentations is largely different from what is associated with quantitative histopathological features. We therefore asked whether plaque proteins carried information on symptomatic presentation beyond histological features. The histological features were partly intercorrelated, with CD68-positive area correlating positively with lipid core (partial Spearman rho = 0.68, FDR = 4.3 × 10^-11^) and intraplaque hemorrhage (partial Spearman rho = 0.64, FDR = 4.8 × 10^-9^), and inversely with calcification (partial Spearman rho = −0.41, FDR = 9.4 × 10^-4^) (**Figure 4A, Supplementary Table 7**). In this limited sample of 88 plaques, only CD68-positive area was significantly associated with symptomatic status after adjustment for age and sex (OR per 1-SD increase = 1.92, 95% CI 1.13– 3.27), while the remaining features, including intraplaque hemorrhage, had wide confidence intervals overlapping the null (**Figure 4B**). In single-feature models, apparent discrimination was modest for histological features, reaching an apparent AUC of 0.71 (95% CI 0.59–0.84) for intraplaque hemorrhage and 0.63 (95% CI 0.51–0.75) for CD68-positive area. Several individual plaque proteins reached higher apparent AUCs, led by FGFBP1 at 0.79 (95% CI 0.69–0.89), CEACAM8 at 0.74 (95% CI 0.63–0.84), and BCL2L15 at 0.73 (95% CI 0.63–0.84) (**Figure 4C**). To compare the incremental information from histology and proteins, we evaluated nested models built on age and sex (**Figure 4D**). Age and sex alone discriminated symptomatic from asymptomatic plaques close to chance (apparent AUC = 0.55, 95% CI 0.44–0.69). Adding intraplaque hemorrhage and CD68-positive area modestly increased discrimination (apparent AUC = 0.66, 95% CI 0.56–0.79), while adding the three leading plaque proteins produced a larger increase (apparent AUC = 0.83, 95% CI 0.74–0.92). The model combining histology and proteins reached an apparent AUC of 0.85 (95% CI 0.77–0.94). Compared with the histopathology model, the protein model increased the apparent AUC by 0.16 (95% CI 0.02–0.31; P = 0.026), while the combined pathology–protein model increased it by 0.19 (95% CI 0.07–0.31; P = 0.003). These patterns were similar in complete-case sensitivity analyses (**Supplementary Figure 2, Supplementary Table 7**).

**Figure 4.**
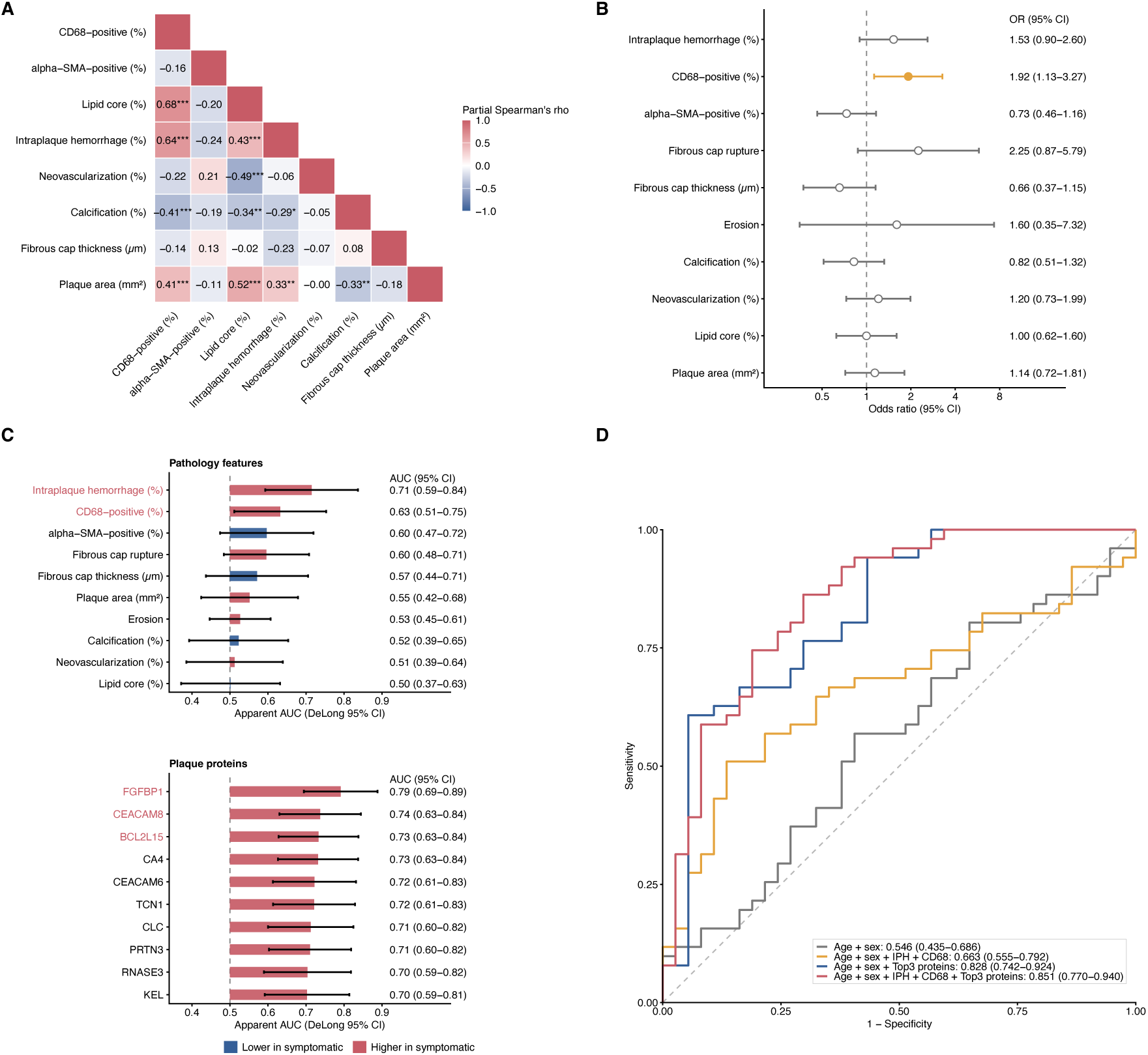
Plaque proteomics adds discriminative information beyond histopathology. **(A)** Pairwise age- and sex-adjusted partial correlations among histopathological plaque features. Values indicate Spearman’s ρ. Asterisks indicate Benjamini-Hochberg-adjusted significance: *FDR < 0.05, **FDR < 0.01, and ***FDR < 0.001. **(B)** Associations between histopathological features and symptomatic plaque presentation. Odds ratios and 95% confidence intervals were estimated using age- and sex-adjusted logistic regression models. Continuous variables are shown per 1-SD increase, and binary variables are shown as yes versus no. Yellow markers indicate P < 0.05. **(C)** Single-feature discrimination of symptomatic plaque presentation by histopathological features and plaque proteins. Bars show apparent AUCs with DeLong 95% confidence intervals. Red labels indicate the top two histopathological features and top three plaque proteins. **(D)** Receiver operating characteristic curves for models including clinical variables alone (age and sex), clinical variables plus selected histopathological features (intraplaque haemorrhage and CD68-positive area), clinical variables plus the top three protein panel (FGFBP1, CEACAM8, and BCL2L15), and the combined model including both pathology and proteins. Values in the legend indicate pooled apparent AUCs with 95% confidence intervals across multiply imputed datasets.

### Single-cell-informed proteomic signatures provide cellular context for plaque biology

Plaque proteomics captures a broad range of disease-associated proteins, but bulk protein measurements do not directly resolve how these signals relate to the cellular compartments present in the lesion. We therefore asked whether Olink-measured plaque proteins could be organized into cell-class-informed proteomic signatures using a human plaque scRNA-seq reference. The integrated public plaque scRNA-seq reference and the plaque snRNA-seq dataset both represented major plaque cell compartments, including macrophages, smooth muscle cells, endothelial cells, fibroblasts, and lymphoid populations (**Figure 5A**). Cell-class representation by symptom status is shown in **Figure 5B**. Using the scRNA-seq reference, cell-class-enriched genes were linked to Olink plaque assays and summarized into proteomic signatures (**Figure 5C**; **Supplementary Table 8**).

**Figure 5.**
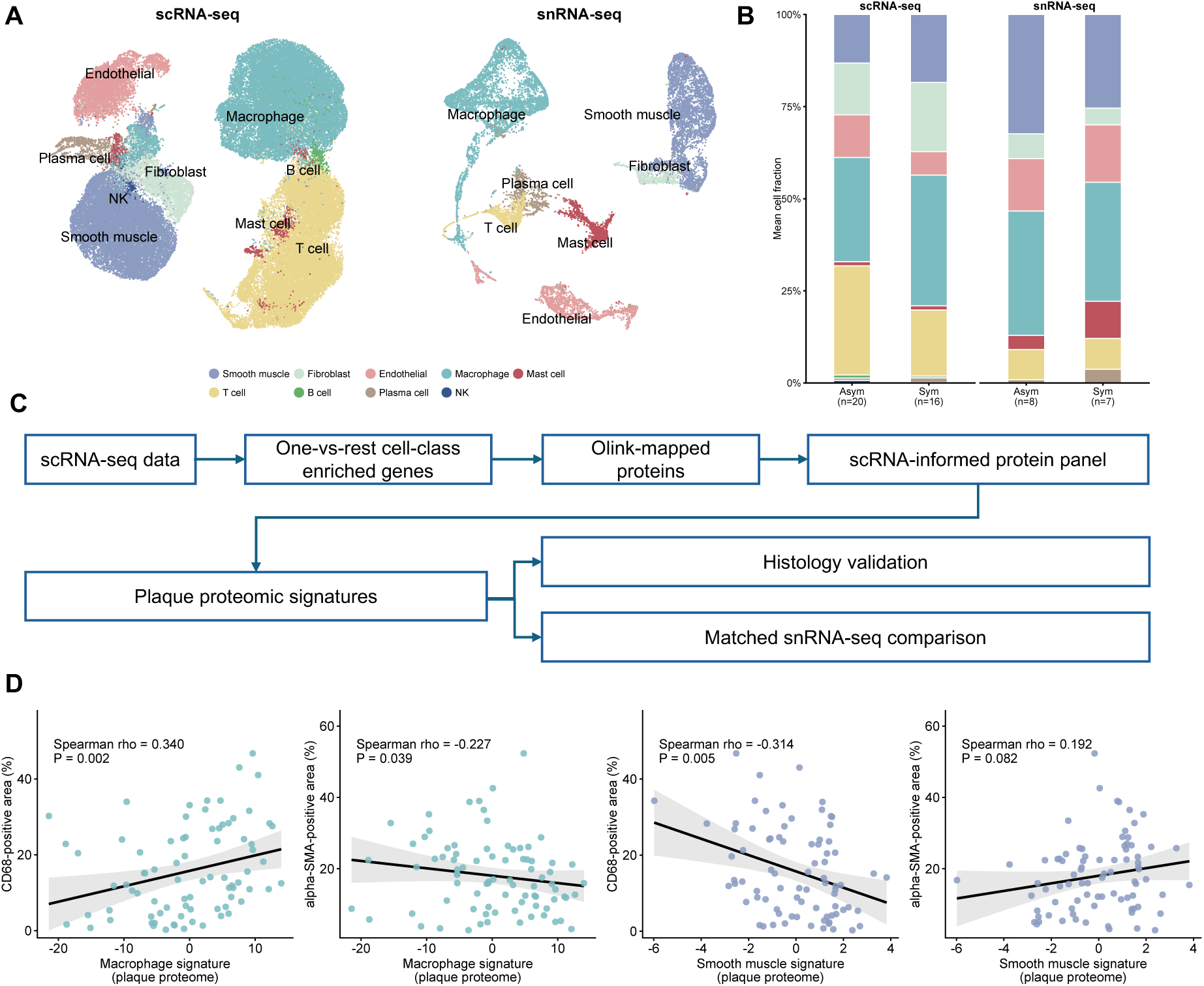
Single-cell-informed signatures provide cellular annotation of the bulk plaque proteome. **(A)** UMAP visualization of broad cell-class annotations in the integrated scRNA-seq reference atlas and the snRNA- seq dataset. **(B)** Cell-class composition by symptom status in the scRNA-seq and snRNA-seq datasets. Bars show group means of sample-level cell or nucleus fractions; n denotes the number of plaques/samples in each group**. (C)** Workflow for signature derivation and validation. One-versus-rest analysis of the scRNA-seq reference atlas was used to identify cell-class enriched genes, which were mapped to Olink assays to define a scRNA-informed protein panel. A sample-level plaque proteomic signature was then calculated and evaluated in the histology measurements and in the matched proteome–snRNA-seq subset. **(D)** Associations between scRNA-informed macrophage and smooth muscle cell (SMC) proteomic signatures and histological CD68-positive or αSMA-positive area in plaque tissue. Spearman correlation coefficients and two-sided P values are shown in the panels. Solid lines indicate linear fits for visualization.

We focused on macrophage and SMC signatures because these compartments represent major inflammatory and structural axes of plaque biology and can be directly compared with CD68-positive and αSMA-positive plaque area. The macrophage signature comprised 127 Olink assays, with PC1 explaining 52.0% of panel variance, and the SMC signature comprised 6 assays, with PC1 explaining 49.6%. The macrophage proteomic signature was positively associated with CD68-positive area (Spearman rho = 0.34, P = 0.002; n = 80), consistent with higher histological macrophage burden, whereas the SMC signature was inversely associated with it (rho = −0.32, P = 0.005; n = 80) (**Figure 5D**). For αSMA-positive area, the pattern reversed. The macrophage signature was inversely associated with αSMA-positive area (rho = −0.23, P = 0.039; n = 83), while the SMC signature showed a weaker positive trend (rho = 0.20, P = 0.082; n = 83) (**Figure 5D**).

The broader cell-class signature screen showed histologically coherent associations for several compartments. The endothelial signature was most strongly associated with neovascularization (rho = 0.43, FDR = 3.5 × 10^-3^), consistent with the role of endothelial cells in plaque angiogenesis, whereas the T cell signature showed a positive trend with the snRNA-seq CD4+ T cell fraction (rho = 0.57, P = 0.026) and was inversely associated with calcification (rho = −0.35, FDR = 2.0 × 10^-2^). Several immune-cell signatures co-varied with neovascularization, consistent with immune infiltration of neovascularized regions (**Supplementary Figure 3; Supplementary Table 9**). Overall, these associations indicate that the cell-class proteomic signatures could uncover the plaque cellular architecture.

### Plasma proteomic changes show partial concordance with matched plaque signals

As a final step, we examined symptom-associated protein differences in matched plasma and then assessed the extent of within-person plaque–plasma correspondence. Across the 2,841 plasma proteins quantified in the same patients, age- and sex-adjusted differential abundance analysis identified a plasma signal led by higher interleukin-6 (IL6; log2FC = 1.40, FDR = 8.8 × 10^-4^) and SERPINA3 (log2FC = 0.37, FDR = 7.7 × 10^-3^), together with lower HPGDS (log2FC = −0.65, FDR = 2.3 × 10^-3^; **Figure 6A, Supplementary Table 10**). When comparing stroke or RAO vs. asymptomatic plaques, there were 10 differentially abundant proteins at FDR < 0.05; IL6 remained elevated (log2FC = 1.58, FDR = 1.4 × 10^-3^), alongside higher glial fibrillary acidic protein (GFAP; log2FC = 2.01, FDR = 1.4 × 10^-3^), CEND1 (log2FC = 1.75, FDR = 4.3 × 10^-3^), and neurofilament light chain (NEFL; log2FC = 1.38, FDR = 8.3 × 10^-3^; **Figure 6B, Supplementary Table 11**). The increases in GFAP and NEFL are consistent with downstream neural injury, whereas the prominent IL6 and SERPINA3 signals probably indicate a systemic inflammatory response associated with symptomatic disease.^32,33^ Reactome enrichment of the plasma contrast showed upregulation of cellular stress-response pathways and downregulation of immunoregulatory lymphoid–non-lymphoid interactions (**Supplementary Table 12**).

**Figure 6.**
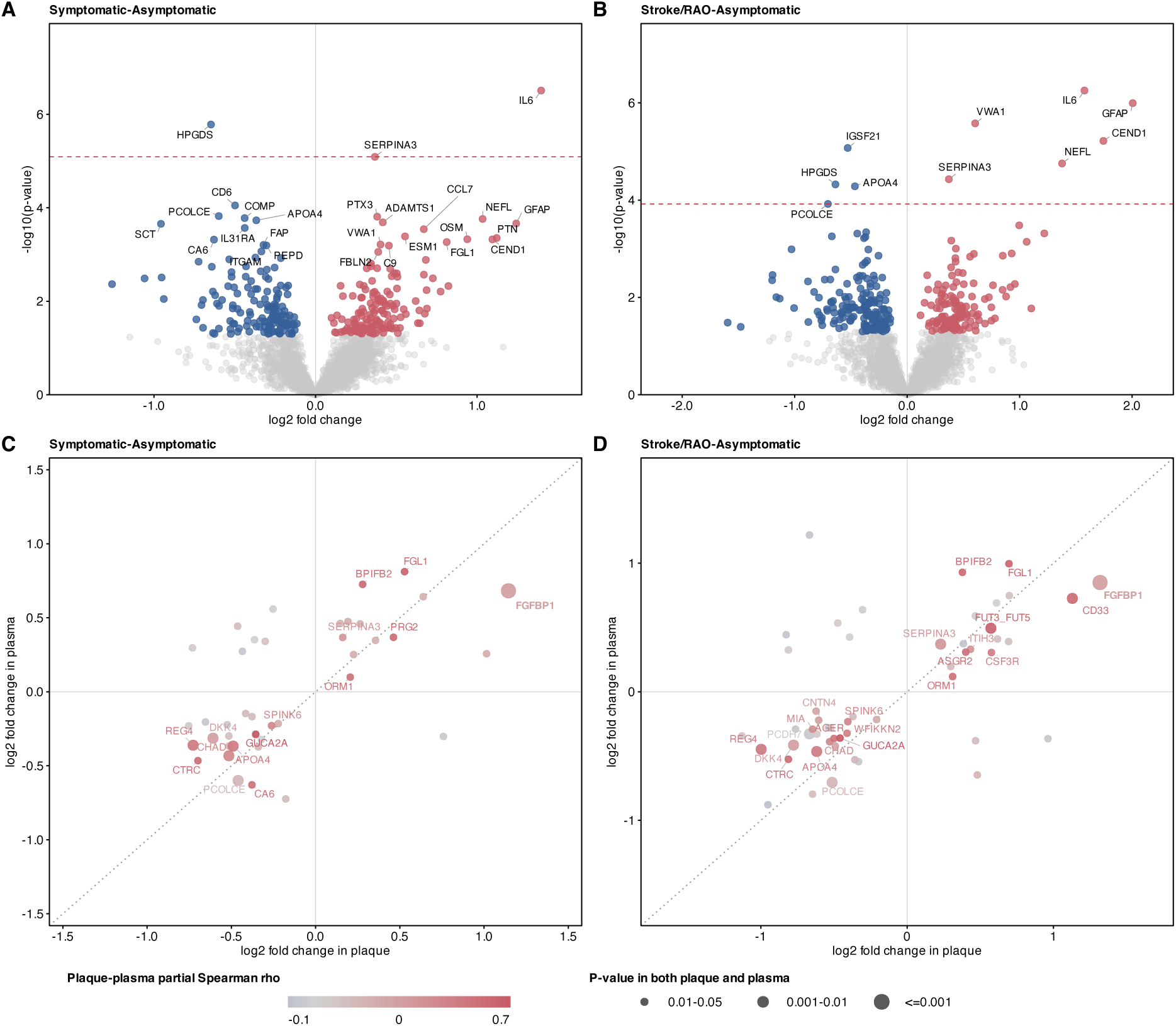
Plasma proteomic signals associated with symptomatic carotid plaque show only partial concordance with matched plaque signals. **(A)** Volcano plot of plasma protein differential abundance for symptomatic versus asymptomatic plaques. The x-axis shows age- and sex-adjusted log2 fold change and the y-axis shows −log10(P value). Proteins with P < 0.05 are coloured red or blue according to positive or negative log2 fold change, respectively; all other proteins are shown in grey. The horizontal dashed line indicates the panel-specific P-value boundary corresponding to BH-adjusted P < 0.05. Labels are shown for proteins with adjusted P < 0.10. **(B)** Volcano plot of plasma protein differential abundance for stroke/RAO versus asymptomatic plaques, displayed as in **(A).** Labels are shown for proteins with adjusted P < 0.05. **(C)** Scatter plot of proteins nominally associated in both plaque and plasma for symptomatic versus asymptomatic plaques. Only proteins with P < 0.05 in both compartments are shown. Axes indicate age- and sex-adjusted plaque and plasma log2 fold changes. Point colour denotes the matched plaque–plasma Spearman correlation coefficient. Point size denotes the P-value tier met in both plaque and plasma (0.01–0.05, 0.001–0.01, or ≤0.001). Labels are shown for proteins with P < 0.01 in both compartments, or for proteins with P < 0.05 in both compartments and plaque–plasma Spearman rho > 0.30. **(D)** Scatter plot of proteins nominally associated in both plaque and plasma for stroke/RAO versus asymptomatic plaques, displayed as in **(C)**.

Among the 2,837 proteins quantified in both plaque and plasma, age- and sex-adjusted partial Spearman correlations were modest overall (median ρ = 0.11), with 367 proteins reaching ρ > 0.30 at FDR < 0.05 (**Supplementary Table 13**). For proteins nominally significant in both plaque and plasma, only a small set changed in the same direction in both compartments (**Figure 6C, 6D, Supplementary Table 14**). FGFBP1 and CD33 were higher in both plaque and plasma, whereas Regenerating Family Member 4 (REG4), apolipoprotein A4 (APOA4), and procollagen C-endopeptidase enhancer (PCOLCE) were lower in both, indicating potential utility for the development of biomarkers of symptomatic disease.

## Discussion

In this paired plaque–plasma proteomic study of patients undergoing carotid endarterectomy, we characterized plaque protein signals of symptomatic atherosclerotic disease across four dimensions of lesion biology. First, comparisons between plaques associated with symptomatic presentations showed enrichment in FGFBP1, as well as for neutrophil degranulation-associated proteins. Second, while plaque proteins were associated with histopathological features of plaque vulnerability, the symptom-associated and histopathology-associated protein sets showed limited overlap. Notably, symptom-associated proteins showed greater apparent discrimination of symptomatic presentation than established histopathological features. Third, we integrated our data with data from scRNA to develop protein signatures of macrophage and SMC content that tracked matched histology, linking the plaque proteome to cellular abundance. Fourth, paired plasma profiling showed that, overall, only a subset of plaque-associated signals are detectable in the circulation and could serve the development of circulating biomarkers.

Our findings refine the molecular characterization of symptomatic carotid stenosis. FGFBP1 was the leading plaque protein in the symptomatic–asymptomatic comparison. As a secreted fibroblast-growth-factor-binding protein, FGFBP1 can mobilize extracellular matrix-bound FGFs and potentiate FGF signaling. Its identification in symptomatic plaques raises the possibility that extracellular growth-factor availability is altered in clinically active lesions and prioritizes FGFBP1 for spatial and functional investigation in human carotid stenosis.^34^ Further stratification by clinical presentation revealed a broader neutrophil-related program in stroke/RAO plaques, including CEACAM8, PRTN3, AZU1, and MMP8, together with enrichment of neutrophil degranulation. This program was supported by the histopathology analyses, in which its leading-edge proteins showed coordinated nominal associations with greater CD68-positive area, intraplaque hemorrhage and lower αSMA-positive area. CXCL8 provided an additional inflammatory link, as it was the strongest protein associated with both CD68-positive area and lipid core and also increased with PVI. Consistent with previous observations, plaque proteomics has linked the plaque inflammation axis to neutrophil-derived proteins,^9^ and human carotid plaques rich in neutrophils show greater macrophage burden, larger lipid cores, reduced smooth muscle and collagen content, increased neovessel density, and higher CXCL8, MMP8, and MMP9, with active MMP8 elevated in unstable plaques.^35–37^

The histopathology analyses further revealed molecular programs related to vascular remodeling, calcification, and composite plaque vulnerability. Neovascularization had the broadest set of FDR-significant associations, with EPHB4 and PKN3 among the leading proteins. EPHB4 regulates postnatal vascular morphogenesis, whereas loss of PKN3 impairs growth-factor-induced microvascular sprouting, indicating that angiogenesis and remodeling programs play a notable role in highly vascularized plaques.^30,31^ Calcification showed a predominantly inverse association profile, consistent with the inversely correlated inflammation and calcification proteomic signatures

previously identified in human carotid plaques.^9^ At the composite level, higher PVI was associated with enrichment of inflammatory response, TNF–NFκB signaling, complement, hypoxia, and IL6– JAK–STAT3 signaling, highlighting inflammatory signaling as a prominent molecular feature of histologically vulnerable plaques. These findings reinforce inflammation as a therapeutically relevant component of plaque vulnerability and provide a rationale for testing whether targeted anti-inflammatory therapy can stabilize high-risk carotid plaques. Plaque proteomics did more than provide molecular readouts of quantitative histopathology. A three-protein model discriminated symptomatic presentation more strongly than an age- and sex-adjusted model incorporating CD68-positive area and intraplaque hemorrhage (apparent AUC, 0.83 versus 0.66), and combining the two reached the highest apparent discrimination (AUC, 0.85). By comparing clinical presentation, multiple quantitative histological features, and plaque protein abundance within the same plaques, we found that the plaque proteome captures both molecular correlates of plaque morphology and additional symptom-associated information not fully represented by measured histopathology.

Single-cell atlases have resolved the cellular diversity of human atherosclerotic plaques, but whether the bulk plaque proteome reflects this cellular composition is less clear. Previous plaque proteomic studies first identified proteins associated with plaque vulnerability and its hallmark pathways before determining likely orgin cell types for each target protein using single-cell expression data.^9,19^ We adopted cell-type-first approach based on TEMPO,^22^ by building protein signatures for each cell class from public human carotid plaque scRNA-seq data and applying said signatures to our proteomic dataset. The strongest pattern was a coordinated increase in macrophage-enriched proteins and decrease in smooth-muscle-cell-enriched proteins with greater CD68 burden. Neovascularization was associated with endothelial and several immune-cell signatures, consistent with immune infiltration of neovascularized regions. These patterns were recovered from bulk tissue, in which cell types are mixed, showing that bulk proteins alone can report part of the cellular landscape of a plaque. As single-cell atlases expand, this approach can be refined from broad cell classes to disease-relevant cell states and used to prioritize targets for spatial and functional validation.

In line with previous studies, same-protein correlations between plaque and plasma were modest overall.^14,18^ Nevertheless, selected lesion-associated proteins can show corresponding circulating signals. Among proteins nominally associated with symptomatic events in both compartments, FGFBP1 was concordantly increased in the broad symptomatic comparison and remained so in stroke/RAO, where CD33 emerged as an additional concordant signal. The plasma findings also extended beyond these concordant proteins: IL6 and SERPINA3 characterized the broad symptomatic comparison, whereas GFAP and NEFL emerged in stroke/RAO. Circulating IL6 has previously been reported to be higher in symptomatic carotid disease, while GFAP and NEFL are established markers of glial and neuroaxonal injury after ischemic stroke.^32,38^ Matched plaque–plasma profiling thus placed the circulating signal in tissue context, revealing a limited plaque-concordant component alongside broader inflammatory and neural-injury signals.

Our study has limitations. First, the study included 88 patients. This limited power for separate comparisons of stroke, RAO, TIA, and amaurosis fugax against asymptomatic plaques, and precluded splitting the data into development and validation sets, so protein selection, model fitting, and performance assessment were conducted in the same dataset. The reported AUCs therefore represent apparent within-study discrimination, and the incremental value of plaque proteins requires confirmation in independent cohorts. The matched single-nucleus subset was likewise small (n = 15), in which the T-cell signature showed nominal positive associations with corresponding T-cell fractions whereas most other signatures were weak or inconsistent. This subset was not sufficient to validate cell-class attribution. Second, all participants, including those classified as asymptomatic, had advanced carotid stenosis selected for endarterectomy, so the findings may not extend to earlier lesions. Third, in symptomatic patients, plaque tissue was obtained at endarterectomy after the ischemic event, at a median of 6 days from symptom onset (documented in 40 of 51 patients; range 1–70 days). Thrombus organization, inflammatory resolution, and remodeling during this interval may have altered the molecular and histopathological state of the culprit plaque relative to the time of symptom onset. In addition, transient symptomatic events such as TIA and amaurosis fugax reply on self-reported symptoms days to weeks prior to presentation, which may reduce the precision of clinical subtyping. This likely diluted the broad symptomatic comparison, in which only one protein reached FDR<0.05, compared with 19 in the stroke/RAO subgroup. Fourth, histopathology was assessed in the most-diseased segment, whereas proteomics was performed in the immediately adjacent plaque segment. Although this design minimized spatial separation, heterogeneity within advanced plaques may have attenuated associations between protein abundance and focal histopathological features. Fifth, the cell-class signatures were derived from public scRNA-seq data. They annotate plaque protein variation rather than measure the cellular source of individual proteins or the abundance of each cell type. Sixth, concordant plaque–plasma changes indicate cross-compartment correspondence but do not establish that the circulating proteins were released from the operated lesion.

In conclusion, deep proteomic profiling identified molecular signatures of symptomatic carotid atherosclerosis that extended beyond conventional histopathology. Neutrophil activation and inflammatory signaling emerged as prominent molecular features of vulnerable and symptomatic plaques. Although plaque and plasma proteomes were largely distinct, selected plaque-concordant proteins warrant further evaluation as potential circulating biomarkers.

## Data Availability

All data produced in the present work are contained in the manuscript

## Acknowledgments

The authors are grateful to all participants of the AtherOMICS Biobank for their participation in and contribution to this study.

---

**Supplementary Figure 1.**
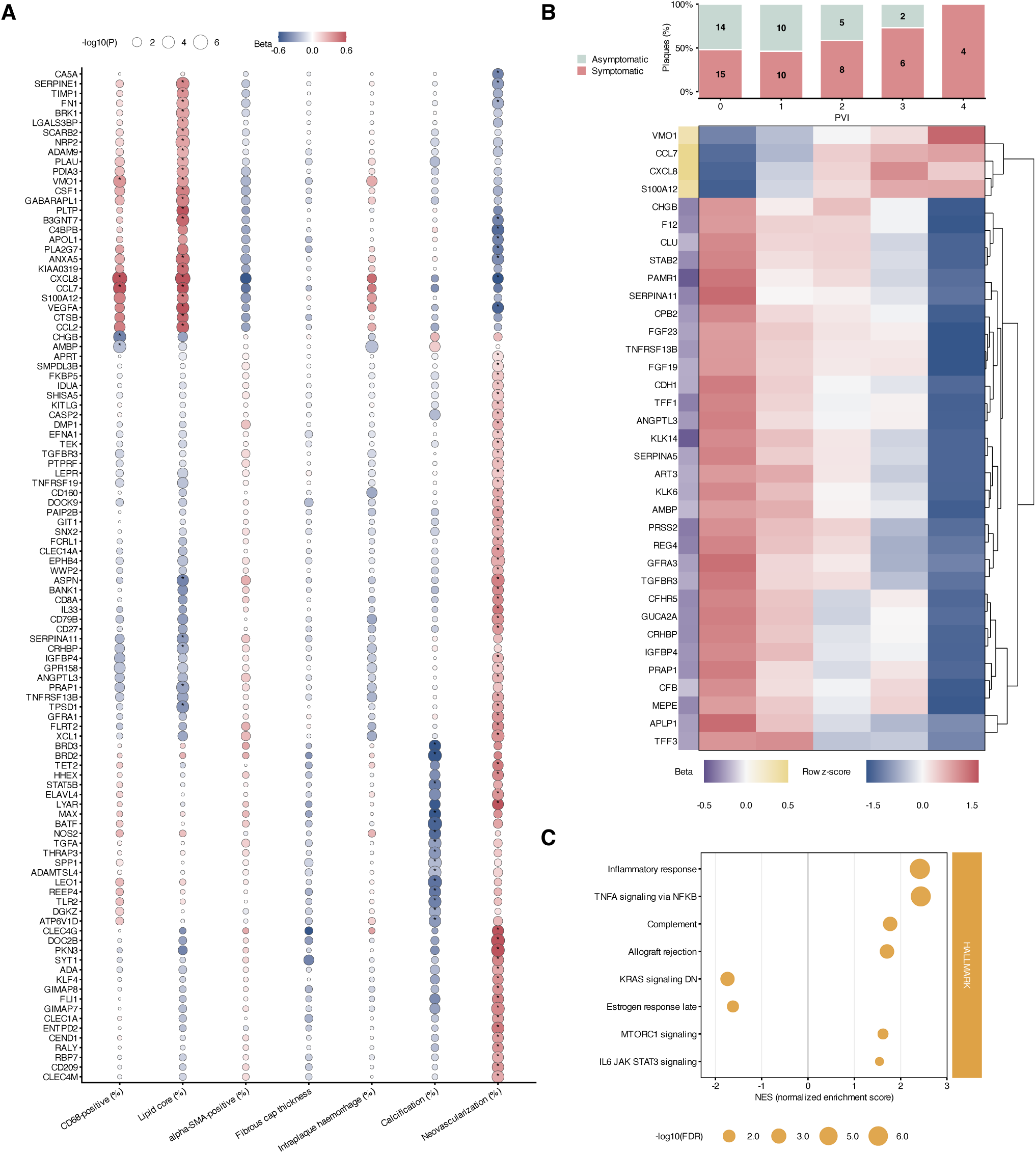
Complete-case analysis of plaque proteomic associations with lesion pathology and vulnerability. Using plaques with complete observed pathology data, plaque protein abundance was analyzed in relation to histopathological features and the plaque vulnerability index (PVI). **(A)** Bubble plot showing the union of plaque proteins significantly associated with individual histopathological features in feature-specific regression analyses. Columns represent CD68-positive area, lipid core, αSMA-positive area, fibrous cap thickness, intraplaque hemorrhage, calcification, and neovascularization. Bubble color indicates the regression coefficient (beta), bubble size indicates -log10(P value), and asterisks denote FDR < 0.05. **(B)** Heatmap of plaque proteins significantly associated with the PVI. Columns represent PVI groups from 0 to 4, and values are shown as row z-scores of age- and sex-adjusted mean protein abundance. The side annotation shows the PVI association coefficient. Stacked bars above the heatmap show the proportions of symptomatic and asymptomatic plaques within each PVI group, with numbers indicating patient counts. **(C)** Hallmark pathway enrichment analysis based on proteins ranked by their PVI association statistics. Bubble position indicates the normalized enrichment score (NES), and bubble size indicates -log10(FDR).

**Supplementary Figure 2.**
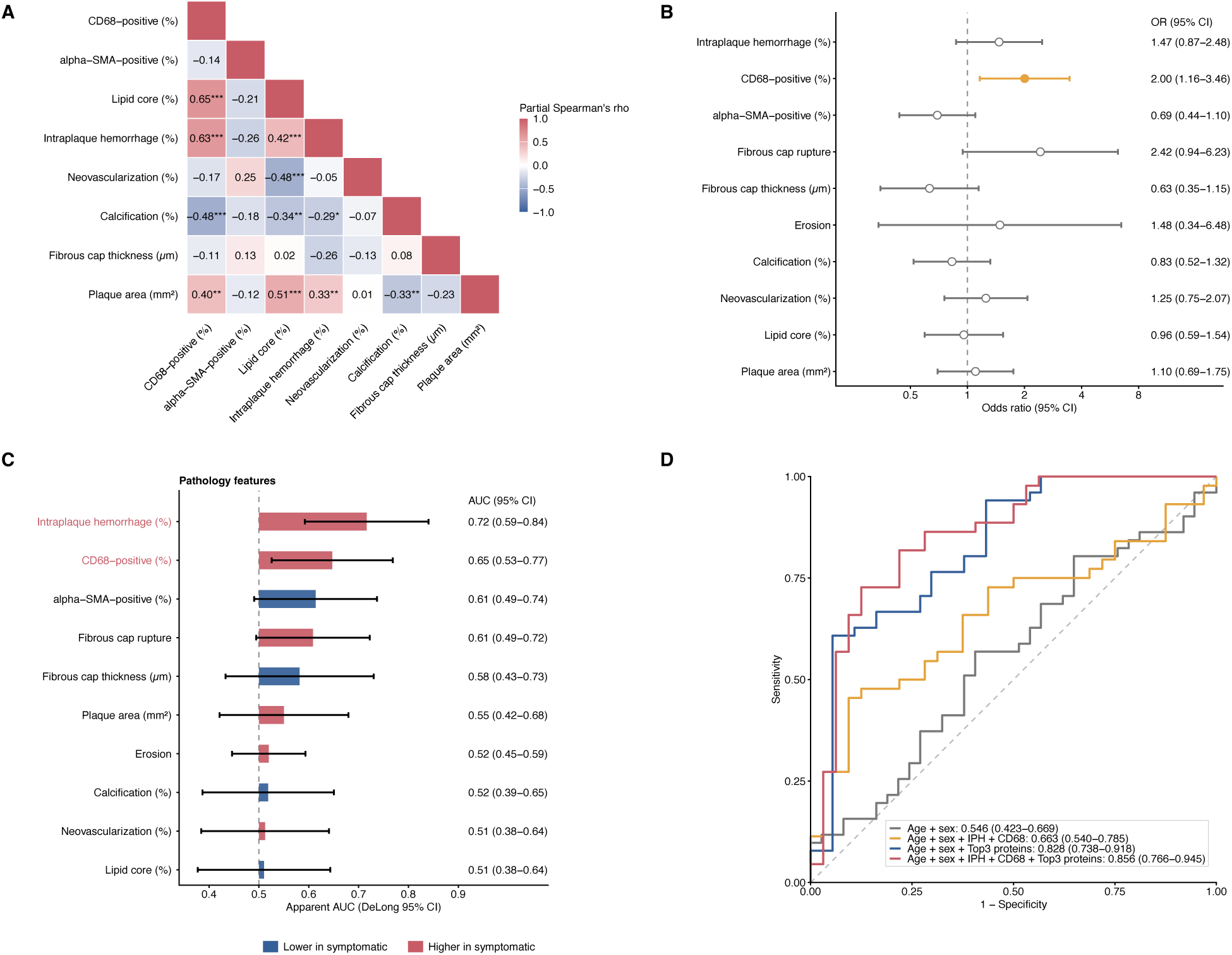
Complete-case analysis of plaque histopathology and proteomics for symptomatic plaque classification. **(A)** Pairwise age- and sex-adjusted partial correlations among histopathological plaque features. Values indicate Spearman’s ρ. Asterisks indicate Benjamini-Hochberg-adjusted significance: *FDR < 0.05, **FDR < 0.01, and ***FDR < 0.001. **(B)** Associations between histopathological features and symptomatic plaque presentation. Odds ratios and 95% confidence intervals were estimated using age- and sex-adjusted logistic regression models. Continuous variables are shown per 1-SD increase, and binary variables are shown as yes versus no. Yellow markers indicate P < 0.05. **(C)** Single-feature discrimination of symptomatic plaque presentation by histopathological features and plaque proteins. Bars show apparent AUCs with DeLong 95% confidence intervals. Red labels indicate the top two histopathological features and top three plaque proteins. **(D)** Receiver operating characteristic curves for models including clinical variables alone (age and sex), clinical variables plus selected histopathological features (intraplaque hemorrhage and CD68-positive area), clinical variables plus the top three protein panel (FGFBP1, CEACAM8, and BCL2L15), and the combined model including both pathology and proteins. Values in the legend indicate apparent AUCs with DeLong 95% confidence intervals.

**Supplementary Figure 3.**
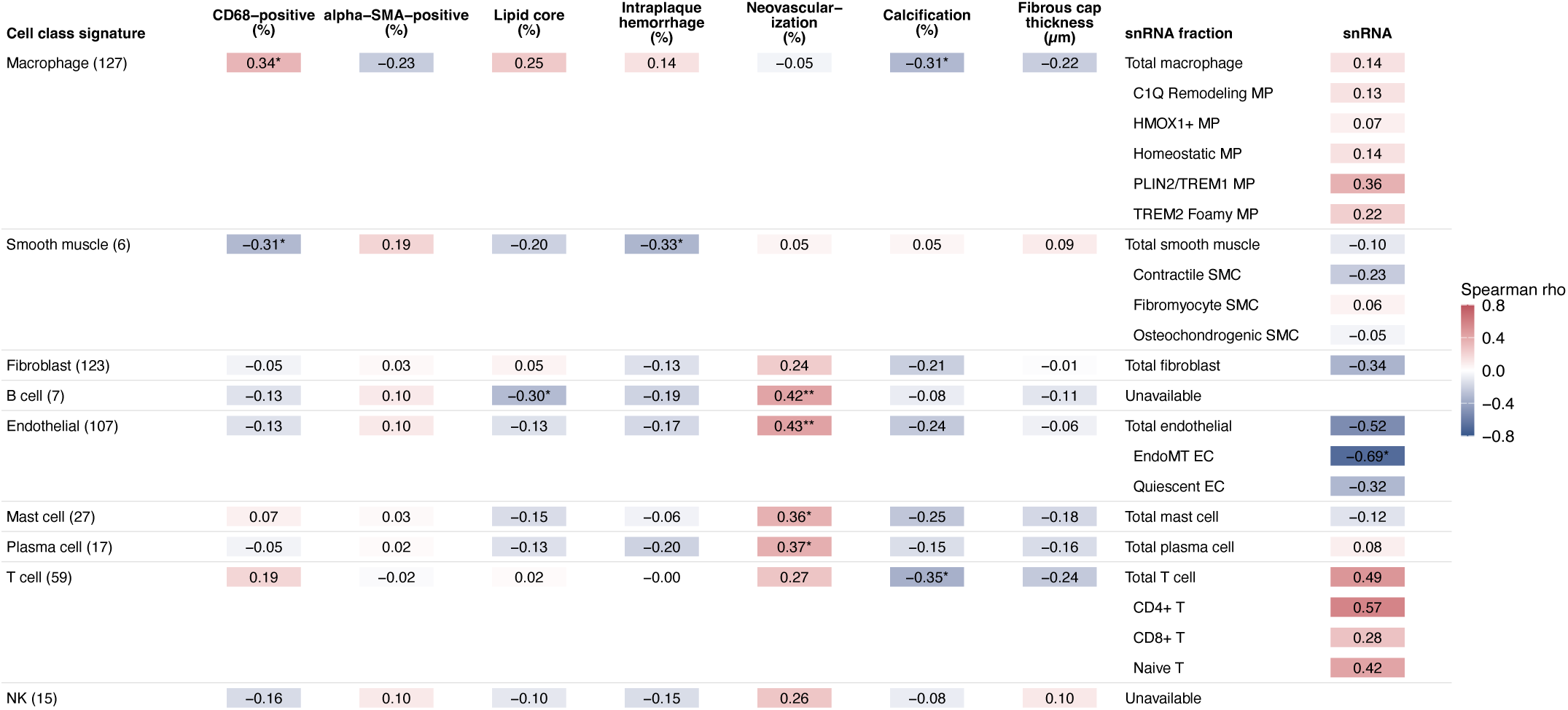
Cell-class plaque proteomic signatures across histology and matched snRNA-seq features.

## References

1. Palaniappan LP, Allen NB, Almarzooq ZI, Anderson CAM, Arora P, Avery CL, Baker-Smith CM, Bansal N, Currie ME, Earlie RS, et al. 2026 Heart Disease and Stroke Statistics: A Report of US and Global Data From the American Heart Association. Circulation. 2026;153:e275–e906. doi: 10.1161/CIR.0000000000001412

2. Global Burden of Cardiovascular Diseases and Risks 2023 Collaborators. Global, Regional, and National Burden of Cardiovascular Diseases and Risk Factors in 204 Countries and Territories, 1990-2023. J Am Coll Cardiol. 2025;86:2167–2243. doi: 10.1016/j.jacc.2025.08.015

3. Barrett KM, Brott TG. Stroke Caused by Extracranial Disease. Circ Res. 2017;120:496–501. doi: 10.1161/CIRCRESAHA.117.310138

4. Falk E, Shah PK, Fuster V. Coronary plaque disruption. Circulation. 1995;92:657–671. doi: 10.1161/01.cir.92.3.657

5. Virmani R, Burke AP, Farb A, Kolodgie FD. Pathology of the vulnerable plaque. J Am Coll Cardiol. 2006;47:C13–18. doi: 10.1016/j.jacc.2005.10.065

6. Carr S, Farb A, Pearce WH, Virmani R, Yao JS. Atherosclerotic plaque rupture in symptomatic carotid artery stenosis. J Vasc Surg. 1996;23:755–765; discussion 765-756. doi: 10.1016/s0741-5214(96)70237-9

7. Hatsukami TS, Ferguson MS, Beach KW, Gordon D, Detmer P, Burns D, Alpers C, Strandness DE, Jr. Carotid plaque morphology and clinical events. Stroke. 1997;28:95–100. doi: 10.1161/01.str.28.1.95

8. Spagnoli LG, Mauriello A, Sangiorgi G, Fratoni S, Bonanno E, Schwartz RS, Piepgras DG, Pistolese R, Ippoliti A, Holmes DR, Jr. Extracranial thrombotically active carotid plaque as a risk factor for ischemic stroke. JAMA. 2004;292:1845–1852. doi: 10.1001/jama.292.15.1845

9. Theofilatos K, Stojkovic S, Hasman M, van der Laan SW, Baig F, Barallobre-Barreiro J, Schmidt LE, Yin S, Yin X, Burnap S, et al. Proteomic Atlas of Atherosclerosis: The Contribution of Proteoglycans to Sex Differences, Plaque Phenotypes, and Outcomes. Circ Res. 2023;133:542–558. doi: 10.1161/CIRCRESAHA.123.322590

10. Langley SR, Willeit K, Didangelos A, Matic LP, Skroblin P, Barallobre-Barreiro J, Lengquist M, Rungger G, Kapustin A, Kedenko L, et al. Extracellular matrix proteomics identifies molecular signature of symptomatic carotid plaques. J Clin Invest. 2017;127:1546–1560. doi: 10.1172/JCI86924

11. Nurmohamed NS, Kraaijenhof JM, Mayr M, Nicholls SJ, Koenig W, Catapano AL, Stroes ESG. Proteomics and lipidomics in atherosclerotic cardiovascular disease risk prediction. Eur Heart J. 2023;44:1594–1607. doi: 10.1093/eurheartj/ehad161

12. Lepedda AJ, Cigliano A, Cherchi GM, Spirito R, Maggioni M, Carta F, Turrini F, Edelstein C, Scanu AM, Formato M. A proteomic approach to differentiate histologically classified stable and unstable plaques from human carotid arteries. Atherosclerosis. 2009;203:112–118. doi: 10.1016/j.atherosclerosis.2008.07.001

13. Lorentzen LG, Yeung K, Eldrup N, Eiberg JP, Sillesen HH, Davies MJ. Proteomic analysis of the extracellular matrix of human atherosclerotic plaques shows marked changes between plaque types. Matrix Biol Plus. 2024;21:100141. doi: 10.1016/j.mbplus.2024.100141

14. Verwer MC, Mekke J, Timmerman N, Waissi F, Boltjes A, Pasterkamp G, de Borst GJ, de Kleijn DPV. Comparison of cardiovascular biomarker expression in extracellular vesicles, plasma and carotid plaque for the prediction of MACE in CEA patients. Sci Rep. 2023;13:1010. doi: 10.1038/s41598-023-27916-6

15. Jin H, Goossens P, Juhasz P, Eijgelaar W, Manca M, Karel JMH, Smirnov E, Sikkink C, Mees BME, Waring O, et al. Integrative multiomics analysis of human atherosclerosis reveals a serum response factor-driven network associated with intraplaque hemorrhage. Clin Transl Med. 2021;11:e458. doi: 10.1002/ctm2.458

16. Matic LP, Jesus Iglesias M, Vesterlund M, Lengquist M, Hong MG, Saieed S, Sanchez-Rivera L, Berg M, Razuvaev A, Kronqvist M, et al. Novel Multiomics Profiling of Human Carotid Atherosclerotic Plaques and Plasma Reveals Biliverdin Reductase B as a Marker of Intraplaque Hemorrhage. JACC Basic Transl Sci. 2018;3:464–480. doi: 10.1016/j.jacbts.2018.04.001

17. Lai Z, Wang C, Liu X, Sun H, Guo Z, Shao J, Li K, Chen J, Wang J, Lei X, et al. Characterization of the proteome of stable and unstable carotid atherosclerotic plaques using data-independent acquisition mass spectrometry. J Transl Med. 2024;22:247. doi: 10.1186/s12967-023-04723-1

18. Sinha A, Sachs N, Kratz E, Pauli J, Steigerwald S, Albrecht V, Nordmann TM, Ugur E, Rodriguez EH, Engl ML, et al. Proteomics reveals spatial and molecular heterogeneities in advanced atherosclerotic carotid artery plaques. Nat Cardiovasc Res. 2026. doi: 10.1038/s44161-026-00827-1

19. Palm KCA, Yin X, Baig F, Theofilatos K, van der Laan SW, de Borst GJ, de Kleijn DPV, Wojta J, Stojkovic S, Mayr M, et al. Proteomic profiling reveals a higher presence of glycolytic enzymes in human atherosclerotic lesions with unfavourable histological characteristics. Cardiovasc Res. 2025;121:1187–1203. doi: 10.1093/cvr/cvaf077

20. Živkoviæ L, Batool R, Louma J, Alabarse PVG, Li Y, Zhang L, Mayrhofer S, Ray A, Antabi MA, Zangas P, et al. Multi-omic profiling of atherosclerosis: protocol and pilot data for the AtherOMICS biobank. Science Advances *In press*. 2025:2025.2006.2017.25329773. doi: 10.1101/2025.06.17.25329773

21. Georgakis MK, van der Laan SW, Asare Y, Mekke JM, Haitjema S, Schoneveld AH, de Jager SCA, Nurmohamed NS, Kroon J, Stroes ESG, et al. Monocyte-Chemoattractant Protein-1 Levels in Human Atherosclerotic Lesions Associate With Plaque Vulnerability. Arterioscler Thromb Vasc Biol. 2021;41:2038–2048. doi: 10.1161/ATVBAHA.121.316091

22. Wolf J, Rasmussen DK, Sun YJ, Vu JT, Wang E, Espinosa C, Bigini F, Chang RT, Montague AA, Tang PH, et al. Liquid-biopsy proteomics combined with AI identifies cellular drivers of eye aging and disease in vivo. Cell. 2023;186:4868–4884 e4812. doi: 10.1016/j.cell.2023.09.012

23. Fernandez DM, Rahman AH, Fernandez NF, Chudnovskiy A, Amir ED, Amadori L, Khan NS, Wong CK, Shamailova R, Hill CA, et al. Single-cell immune landscape of human atherosclerotic plaques. Nat Med. 2019;25:1576–1588. doi: 10.1038/s41591-019-0590-4

24. Pan H, Xue C, Auerbach BJ, Fan J, Bashore AC, Cui J, Yang DY, Trignano SB, Liu W, Shi J, et al. Single-Cell Genomics Reveals a Novel Cell State During Smooth Muscle Cell Phenotypic Switching and Potential Therapeutic Targets for Atherosclerosis in Mouse and Human. Circulation. 2020;142:2060–2075. doi: 10.1161/CIRCULATIONAHA.120.048378

25. Bashore AC, Yan H, Xue C, Zhu LY, Kim E, Mawson T, Coronel J, Chung A, Sachs N, Ho S, et al. High-Dimensional Single-Cell Multimodal Landscape of Human Carotid Atherosclerosis. Arterioscler Thromb Vasc Biol. 2024;44:930–945. doi: 10.1161/ATVBAHA.123.320524

26. Mocci G, Sukhavasi K, Ord T, Bankier S, Singha P, Arasu UT, Agbabiaje OO, Makinen P, Ma L, Hodonsky CJ, et al. Single-Cell Gene-Regulatory Networks of Advanced Symptomatic Atherosclerosis. Circ Res. 2024;134:1405–1423. doi: 10.1161/CIRCRESAHA.123.323184

27. Korsunsky I, Millard N, Fan J, Slowikowski K, Zhang F, Wei K, Baglaenko Y, Brenner M, Loh PR, Raychaudhuri S. Fast, sensitive and accurate integration of single-cell data with Harmony. Nat Methods. 2019;16:1289–1296. doi: 10.1038/s41592-019-0619-0

28. Traeuble K, Munz M, Pauli J, Sachs N, Vafadarnejad E, Carrillo-Roa T, Maegdefessel L, Kastner P, Heinig M. Integrated single-cell atlas of human atherosclerotic plaques. Nat Commun. 2025;16:8255. doi: 10.1038/s41467-025-63202-x

29. Hou Y, Huttenlocher A. Advancing chemokine research: the molecular function of CXCL8. J Clin Invest. 2024;134. doi: 10.1172/JCI180984

30. Yang D, Jin C, Ma H, Huang M, Shi GP, Wang J, Xiang M. EphrinB2/EphB4 pathway in postnatal angiogenesis: a potential therapeutic target for ischemic cardiovascular disease. Angiogenesis. 2016;19:297–309. doi: 10.1007/s10456-016-9514-9

31. Mukai H, Muramatsu A, Mashud R, Kubouchi K, Tsujimoto S, Hongu T, Kanaho Y, Tsubaki M, Nishida S, Shioi G, et al. PKN3 is the major regulator of angiogenesis and tumor metastasis in mice. Sci Rep. 2016;6:18979. doi: 10.1038/srep18979

32. Kowalski RG, Ledreux A, Violette JE, Neumann RT, Ornelas D, Yu X, Griffiths SG, Lewis S, Nash P, Monte AA, et al. Rapid Activation of Neuroinflammation in Stroke: Plasma and Extracellular Vesicles Obtained on a Mobile Stroke Unit. Stroke. 2023;54:e52–e57. doi: 10.1161/STROKEAHA.122.041422

33. Tiedt S, Duering M, Barro C, Kaya AG, Boeck J, Bode FJ, Klein M, Dorn F, Gesierich B, Kellert L, et al. Serum neurofilament light: A biomarker of neuroaxonal injury after ischemic stroke. Neurology. 2018;91:e1338–e1347. doi: 10.1212/WNL.0000000000006282

34. Tassi E, Al-Attar A, Aigner A, Swift MR, McDonnell K, Karavanov A, Wellstein A. Enhancement of fibroblast growth factor (FGF) activity by an FGF-binding protein. J Biol Chem. 2001;276:40247–40253. doi: 10.1074/jbc.M104933200

35. Ionita MG, van den Borne P, Catanzariti LM, Moll FL, de Vries JP, Pasterkamp G, Vink A, de Kleijn DP. High neutrophil numbers in human carotid atherosclerotic plaques are associated with characteristics of rupture-prone lesions. Arterioscler Thromb Vasc Biol. 2010;30:1842–1848. doi: 10.1161/ATVBAHA.110.209296

36. Molloy KJ, Thompson MM, Jones JL, Schwalbe EC, Bell PR, Naylor AR, Loftus IM. Unstable carotid plaques exhibit raised matrix metalloproteinase-8 activity. Circulation. 2004;110:337–343. doi: 10.1161/01.CIR.0000135588.65188.14

37. Peeters W, Moll FL, Vink A, van der Spek PJ, de Kleijn DP, de Vries JP, Verheijen JH, Newby AC, Pasterkamp G. Collagenase matrix metalloproteinase-8 expressed in atherosclerotic carotid plaques is associated with systemic cardiovascular outcome. Eur Heart J. 2011;32:2314–2325. doi: 10.1093/eurheartj/ehq517

38. Koutouzis M, Rallidis LS, Peros G, Nomikos A, Tzavara V, Barbatis C, Andrikopoulos V, Vassiliou J, Kyriakides ZS. Serum interleukin-6 is elevated in symptomatic carotid bifurcation disease. Acta Neurol Scand. 2009;119:119–125. doi: 10.1111/j.1600-0404.2008.01068.x

